# Predicting coronary artery abnormalities in Kawasaki disease: Model development and external validation

**DOI:** 10.1101/2025.08.11.25333461

**Authors:** Qianzhi Wang, Yuya Kimura, Junna Oba, Tetsuo Ishikawa, Takuma Ohnishi, Shogo Akahoshi, Kazuki Iio, Yoshihiko Morikawa, Kazuhiro Sakurada, Tohru Kobayashi, Masaru Miura

## Abstract

**Background:** Kawasaki disease (KD) is an acute, pediatric vasculitis associated with coronary artery abnormality (CAA) development. Echocardiography at month 1 post-diagnosis remains the standard for CAA surveillance despite limitations, including patient distress and increased healthcare burden. With declining CAA incidence due to improved treatment, the need for routine follow-up imaging is being reconsidered. This study aimed to develop and externally validate models for predicting CAA development and guide the need for echocardiography.

**Methods:** This study used two prospective multicenter Japanese registries: PEACOCK for model development and internal validation, and Post-RAISE for external validation. The primary outcome was CAA at the month 1 follow-up, defined as a maximum coronary artery Z score (Zmax) ≥ 2. Twenty-nine clinical, laboratory, echocardiographic, and treatment-related variables obtained within one week of diagnosis were selected as predictors. The models included simple models using the previous Zmax as a single predictor, logistic regression models, and machine learning models (LightGBM and XGBoost). Their discrimination, calibration, and clinical utility were assessed.

**Results:** After excluding patients without outcome data, 4,973 and 2,438 patients from PEACOCK and Post-RAISE, respectively, were included. The CAA incidence at month 1 was 5.5% and 6.8% for the respective group. For external validation, a simple model using the Zmax at week 1 produced an area under the curve of 0.79, which failed to improve by more than 0.02 after other variables were added or more complex models were used. Even the best-performing models with a highly sensitive threshold failed to reduce the need for echocardiography at month 1 by more than 30% while maintaining the number of undiagnosed CAA cases to less than ten. The predictive performance declined considerably when the Zmax was omitted from the multivariable models.

**Conclusions:** The Zmax at week 1 was the strongest predictor of CAA at month 1 post-diagnosis. Even advanced models incorporating additional variables failed to achieve a clinically acceptable trade-off between reducing the need for echocardiography and reducing the number of undiagnosed CAA cases. Until superior predictors are identified, echocardiography at month 1 should remain the standard practice.

**Clinical Perspective:** *What Is New?:*

- The maximum Z score on echocardiography one week after diagnosis was the strongest of 29 variables for predicting coronary artery abnormalities (CAA) in patients with Kawasaki disease.
- Even the most sensitive models had a suboptimal ability to predict CAA development and reduce the need for imaging studies, suggesting they have limited utility in clinical decision-making.

*What Are the Clinical Implications?:*

- Until more accurate predictors are found or imaging strategies are optimized, performing echocardiography at one-month follow-up should remain the standard of care.

## INTRODUCTION

Kawasaki disease (KD) is an acute, systemic vasculitis in children that can be complicated by coronary artery abnormalities (CAA), which in turn can lead to myocardial infarction, making their early detection and intervention of paramount importance.

Echocardiography is the standard method of assessing for CAA because it is non-invasive and repeatable. However, it has several limitations. Patients’ movements and crying can degrade image quality and may require the patients to be sedated. The small diameter of coronary arteries in children demands considerable technical expertise and time to conduct the procedure. Avoiding unnecessary testing is desirable to minimize healthcare costs and lessen the burden on patients.

Progress in treatment has reduced the CAA incidence to less than 5%.^1,2^ However, the appropriate duration of echocardiographic follow-up for patients without CAA remains uncertain. The 2020 Japanese guidelines recommend follow-up echocardiography for all patients for up to five years after KD onset while two major, Western guidelines recommend routine echocardiography for at least four to six.^3–6^ Some studies suggested that a shorter follow-up period may suffice although this suggestion awaits verification by studies with a larger cohort.^7,8^

Recent studies have investigated the use of machine learning to predict CAA as means of facilitating decision-making. These models have demonstrated promising performance; a systematic review reported a pooled concordance index of 0.79 for CAA prediction.^9^ Nevertheless, the current evidence is limited by small sample sizes, inadequate handling of missing data, and a lack of external validation.^10^ In particular, it remains unclear whether machine learning models offer additional predictive value beyond simpler predictors, such as early echocardiographic Z scores, for determining the need for follow-up echocardiography at month 1.

This study aimed to develop and externally validate models for predicting CAA at month 1 using data obtained within one week of KD diagnosis. A high-performing model may identify patients for whom a follow-up at month 1 is unnecessary, thereby avoiding placing a physical and psychological burden on those who do not require the procedure, easing hospital staffing demands, and reducing the overall healthcare cost. The utility of machine learning algorithms was also compared with that of conventional methods.

## METHODS

This study complies with the Declaration of Helsinki and was approved by the institutional review board of Tokyo Metropolitan Children’s Medical Center (No. 2024b-12). Informed consent was obtained by the opt-out method. The study adhered to the TRIPOD+AI reporting framework.^11^ Dr Wang has full access to all the data in the study and takes responsibility for its integrity and analysis. Due to patient privacy and IRB restrictions, the data underlying this study are not available for sharing. Aggregate results are provided in the article and Data Supplement.

### Data description

The cohorts of two previous, prospective studies, the Prospective Study on Efficacy of Acute Treatment in a Multicenter Cohort of Children with Kawasaki Disease (PEACOCK) and the Prospective Observational study on STRAtified treatment with Immunoglobulin plus Steroid Efficacy for Kawasaki disease (Post-RAISE), were used respectively for model development with internal validation and external validation. Both studies contain clinical, laboratory, imaging, and treatment data obtained at more than 30 hospitals, including secondary care hospitals, pediatric tertiary centers, and university hospitals, which were used to evaluate the efficacy and safety of treatments for KD. PEACOCK (UMIN000024937) is an ongoing registry designed to assess acute treatments for KD and contains data on 5,466 children who received a diagnosis of KD between July 30, 2016, and May 4, 2024 at any of the 30 participating hospitals. The findings of interim analyses were previously reported.^12^ Post-RAISE (UMIN000007133) prospectively collected comparable data with a focus on the efficacy and safety of adding prednisolone to intravenous immunoglobulin (IVIG) and aspirin. The cohort of the study included 2,628 children treated between July 1, 2012, and June 30, 2015 at 34 hospitals. The details of the cohort and the findings of the study have been published.^2^

In both cohorts, baseline clinical characteristics, laboratory values, and echocardiographic data were collected at diagnosis (days 0–1). The standard therapy consisted of IVIG 2 g/kg and oral aspirin 30 mg/kg/day until defervescence followed by aspirin 5 mg/kg/day for two months. Patients in either cohort with a Kobayashi score ≥ 5 received prednisolone as an optional adjunctive therapy. In the PEACOCK cohort, patients with a Kobayashi score ≥ 5 and total bilirubin ≥ 1 mg/dL were eligible for optional intravenous methylprednisolone pulse therapy (IVMP) or cyclosporine in addition to prednisolone. Patients with a persistent or recurrent fever 24 hours after the initial therapy were defined as non-responders and received additional treatments. The same assessment protocol was used for both cohorts. Additional laboratory tests were performed on days 1–3 and week 1 post-diagnosis (days 4–9). Follow-up echocardiography was conducted at week 1 (days 4–9), week 2 (days 10–18 if still hospitalized), and month 1 (days 20–50) post-diagnosis.

### Data preparation

Patients who received either prednisolone or IVMP were categorized as receiving prednisolone because both drugs belong to the same steroid class, and IVMP has not been shown to impact outcomes differently.^12^ Forty high-risk patients in the PEACOCK cohort received cyclosporine as the initial adjunctive therapy; however, this information was not included in our analysis because these patients comprised fewer than 1% of the PEACOCK cohort. All other, relevant variables and outcomes were collected uniformly from both cohorts, thereby obviating the need to harmonize the data.

### Outcome

The outcome was the occurrence of CAA at month 1 as defined by Zmax ≥ 2. This cutoff reflects the definition of CAA and aligns with the AHA recommendation that waives further echocardiographic follow-up if the Zmax is initially < 2.5 and remains < 2 at 4–6 weeks.^4^ At each hospital, trained pediatricians or echocardiographers, neither of whom were blinded to the patient information, including previous echocardiographic findings, measured the raw coronary artery diameter. The Z scores were calculated using reference values derived from a Japanese cohort.^13^ The Zmax was defined as the larger of the Z scores of the proximal right coronary artery (RCA) and the proximal left anterior descending artery (LAD).^14^ Z scores for the left main coronary artery and the left circumflex artery were excluded owing to anatomical variations and technical limitations of visualization.^7,15^ Patients without a Zmax measurement at month 1 were excluded.

### Predictors

Data from both cohorts were used. The PEACOCK dataset, which was the larger of the two, was used for model development. Sample size was calculated using the method described by Riley et al. with the “pmsampsize” package in R.^16,17^ With 4,973 patients and a 5.5% event rate for CAA development, the model was capable of including up to 29 predictors, assuming it would explain 15% of the outcome variability. Although this estimate was originally developed for traditional, statistical models, it was applied to the machine learning models in our study due to the lack of a feasible method for sample size calculation.

The 29 predictors comprised three patient characteristics: age^1,18–22^, sex^18^, and days to diagnosis^23–25^; nine laboratory indices: neutrophils^1,21,23,24,26^, hemoglobin^18,25,26^, platelets^1,18,21,25,26^, serum sodium^1^, aspartate transaminase^1^, alanine transaminase^25^, albumin^18,19^, total bilirubin^12^, and C-reactive protein (CRP)^1,20,21,25^; three echocardiographic measurements: the Z scores of the RCA and the LAD and the Zmax^20,21^; and two treatment-related variables: days to treatment initiation^1,18,19^ and adjunctive prednisolone use as initial therapy^2,27^. Since this study aimed to create a model for predicting CAA to determine the need for further echocardiographic follow-up rather than to guide treatment at the time of diagnosis, predictors measured within one week were used. Echocardiographic findings at week 2 post-diagnosis were excluded because the test was performed selectively for refractory patients (36.1% in PEACOCK and 41.5% in Post-RAISE) and was considered less useful for early prediction. Table S1 lists the variables and their definitions, types, units, measurement timing, and missingness.

### Analysis

The PEACOCK was used for model development and internal evaluation while the Post-RAISE was used for external validation. Two datasets created from each registry were used to predict CAA occurrence at month 1; one contained data obtained at diagnosis, and the other contained data up to week 1 post-diagnosis. Three types of analysis, namely, the (i) main analysis, (ii) sensitivity analysis, and (iii) exploratory analysis, were performed for each dataset for a total of 12 analyses. Table 1 summarizes the model selection, handling of missing data, and dataset modifications in conjunction with the explanation provided below.

**Table 1.** Datasets and the number of records per analytical method.

|  | Handling of missingness | Model development and<br>internal validation | External validation | Exploratory evaluation |
| --- | --- | --- | --- | --- |
| **Using data obtained at diagnosis only** |  |  |  |  |
| Main analysis <sup>a</sup> | Patients with a missing Zmax at diagnosis were excluded | PEACOCK<br>n = 4,742 (95.4%) | Post-RAISE<br>n = 2,173 (89.1%) |  |
| Sensitivity analysis <sup>b</sup> | Missing data were imputed | PEACOCK<br>n = 4,973 (100%) | Post-RAISE<br>n = 2,438 (100%) |  |
| Exploratory analysis <sup>c</sup> | No exclusion or imputation of missing data | PEACOCK<br>n = 4,973 (100%) | Post-RAISE<br>n = 2,438 (100%) | 1. Post-RAISE (Zmax available), n = 2,173 (89.1%)<br>2. Post-RAISE (Zmax missing), n = 265 (10.9%) |
| **Using data obtained at both diagnosis and one week after** |  |  |  |  |
| Main analysis <sup>a</sup> | Patients without Zmax at week 1 were excluded | PEACOCK<br>n = 4,686 (94.2%) | Post-RAISE<br>n = 2,188 (89.7%) |  |
| Sensitivity analysis <sup>b</sup> | Missing data were imputed | PEACOCK<br>n = 4,973 (100%) | Post-RAISE<br>n = 2,438 (100%) |  |
| Exploratory analysis <sup>c</sup> | No exclusion or imputation of missing data | PEACOCK<br>n = 4,973 (100%) | Post-RAISE<br>n = 2,438 (100%) | 1. Post-RAISE (Zmax at week 1 available), n = 2,173 (89.1%)<br>2. Post-RAISE (Zmax at week 1 missing, but Zmax at diagnosis available), n = 198 (8.1%)<br>3. Post-RAISE (All Zmax missing), n = 52 (2.1%) |

<sup>a</sup>The models used for the main analysis included the raw Zmax model (Zmax as the sole predictor), the LogReg with Zmax model (the Zmax with logistic regression), the LightGBM model, and the XGBoost model.
<sup>b</sup>The models for the sensitivity analysis included the multivariate logistic regression model in addition to the four models used in the main analysis.
<sup>c</sup>Two machine learning models, the LightGBM model and the XGBoost model, were used for the exploratory analysis.

#### (i) Main analysis

Patients without a Zmax value for each time point were excluded. Four models were developed and evaluated. The first model was the raw Zmax model, which used the Zmax as the sole predictor because of its simplicity and good predictive ability as previously reported.^7,14^ The second was LogReg with the Zmax model, which used the Zmax with logistic regression. The third and fourth models used all the selected predictors with the LightGBM and XGBoost machine learning models, respectively. Since these machine learning models inherently accommodated missing data, imputation was unnecessary.

#### (ii) Sensitivity analysis

In the sensitivity analysis, a missing value for any of the predictors was imputed using the MissForest algorithm, a non-parametric, iterative method based on random forests.^28^ This algorithm can handle both continuous and categorical variables and has been shown to outperform traditional imputation methods, especially when applied to mixed-type datasets. First, a random forest model was trained using the existing data from the development dataset (PEACOCK). Missing entries were iteratively predicted and updated until convergence. After fitting the model to the PEACOCK dataset, the trained model was applied to impute missing values in the Post-RAISE external validation dataset. Single imputation was selected because it performs comparably with multiple imputation and is easy to use in clinical practice.^29^ Imputation was performed separately for each dataset to avoid data leakage. Five models were developed with the imputed datasets, including the four described in the main analysis and a multivariate logistic regression model using all the selected predictors.

#### (iii) Exploratory analysis

The exploratory analysis used all the available data from PEACOCK without imputation or exclusion to train two machine learning models: the LightGBM model and the XGBoost model. The models were then evaluated using three subsets of the Post-RAISE dataset: (1) the full dataset, (2) a subset with the available Zmax values, and (3) a subset with missing Zmax values.

Appropriate models were selected for each analysis, including the (1) raw Zmax model, which used the untransformed Zmax as a single predictor (the raw Zmax model); (2) LogReg with Zmax model, a logistic regression model using the Zmax only; (3) multivariate LogReg model; (4) LightGBM model; and (5) XGBoost model. Logistic regression was chosen for its interpretability while LightGBM and XGBoost were chosen for their ability to handle missing data without imputation and to model non-linear relationships. The predictors were standardized for the logistic regression models. Otherwise, no rescaling, transformation, or standardization was performed.

The raw Zmax models explored the predictive performance of various Zmax cutoffs at each time point (at diagnosis and week 1). Discrimination was evaluated using the receiver operating characteristic (ROC) curve and the area under the curve (AUC). A cutoff that maximized specificity while maintaining a sensitivity of at least 0.95 in the PEACOCK dataset was applied to the Post-RAISE dataset. This threshold was determined by experts to minimize false negatives because misclassifying children with CAA had the potential to delay follow-up. Using this threshold, performance metrics, including sensitivity, specificity, positive predictive value (PPV), and negative predictive value (NPV), were calculated for the Post-RAISE dataset. False negative cases, that is, cases where CAA was incorrectly predicted not to occur, were also analyzed.

Logistic regression models were trained, and the AUC was calculated using stratified five-fold cross-validation on the PEACOCK dataset for internal validation. Then, the models were retrained on the entire PEACOCK dataset and applied to the Post-RAISE dataset to assess discrimination (ROC and AUC), calibration (calibration plot and Brier score), and clinical utility (decision curve analysis) for external validation. The cutoff values were chosen and the false negative cases identified using the same procedure as for the raw Zmax models.

To address class imbalance and improve the performance of the LightGBM and XGBoost models, hyperparameter tuning and undersampling with bagging (UnderBagging) were performed (see Supplementary Methods).^30^ Internal and external validation, the cutoff selection, and identification of false negative cases followed the same procedure as for the logistic models.

## RESULTS

In total, 493 and 190 children from the PEACOCK and Post-RAISE cohort, respectively, were excluded owing to missing outcome data, specifically the echocardiographic findings at month 1. Tables S2 and S3 compare the baseline characteristics of the included and excluded patients. Table 2 summarizes the characteristics of the 4,973 children from the PEACOCK and 2,438 children from the Post-RAISE who were included in our analysis. Children with CAA were younger and had a higher median Kobayashi score than those without CAA. They also had a higher, median values for aminotransferases and CRP; dilation of the coronary arteries on echocardiography prior to the month 1 assessment; and a greater likelihood of receiving adjunctive prednisolone as part of their initial therapy and of requiring second-line treatment for refractory disease. Notably, a lower percentage of the PEACOCK cohort received adjunctive steroids while a higher proportion required second-line therapy. The background characteristics of the two cohorts, including the CAA incidence, which was 5.5% in PEACOCK and 6.8% in Post-RAISE, were generally comparable.

**Table 2.**
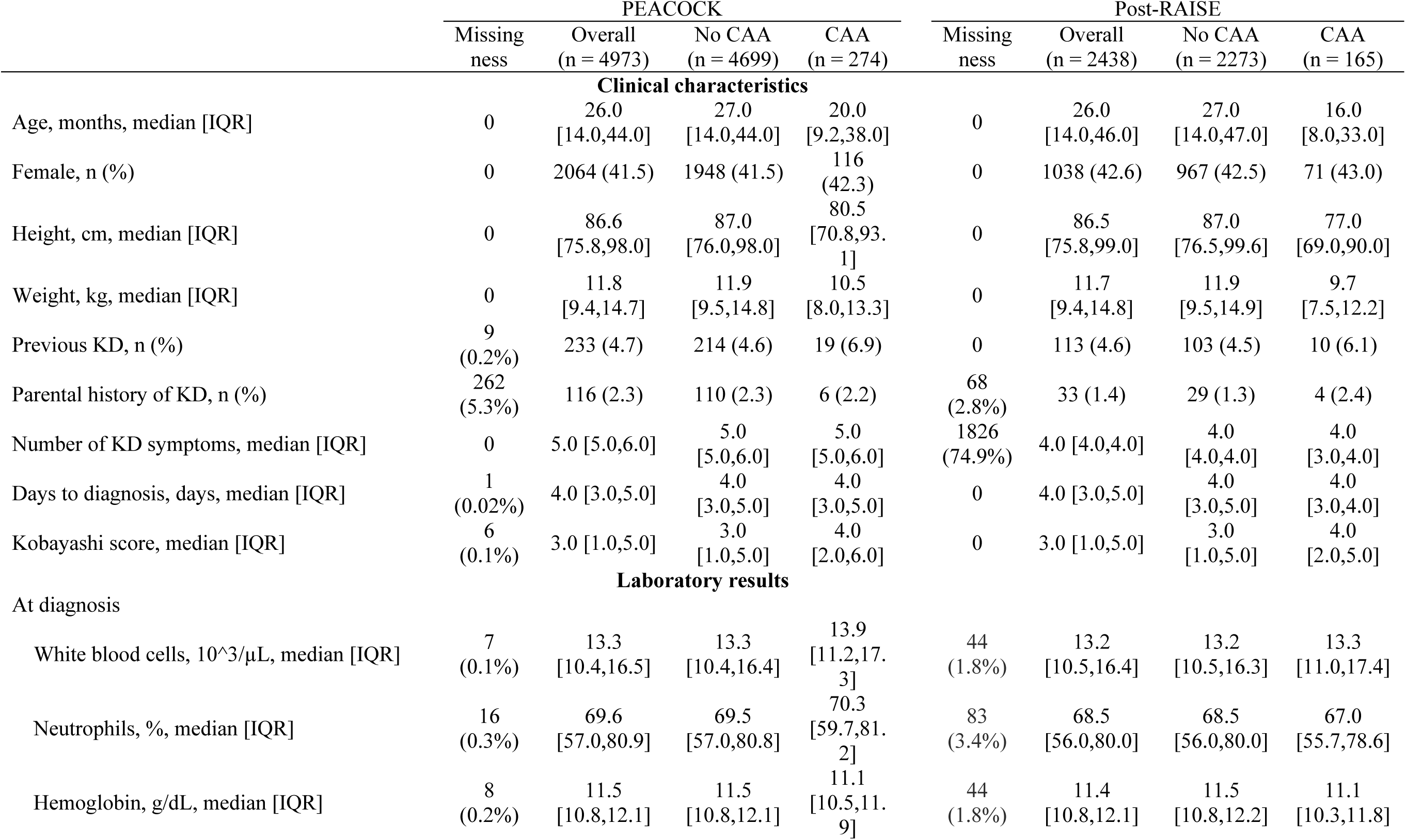
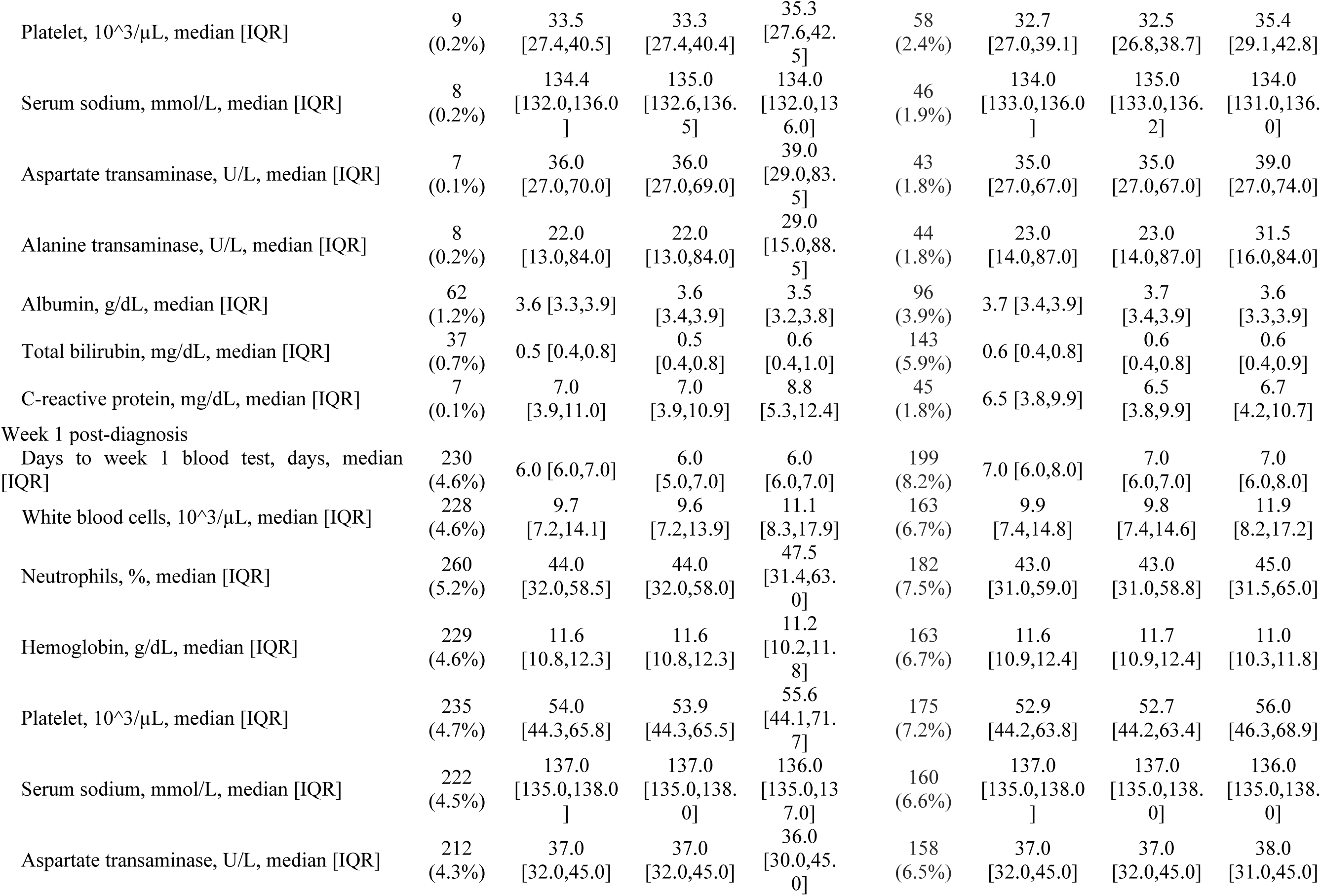
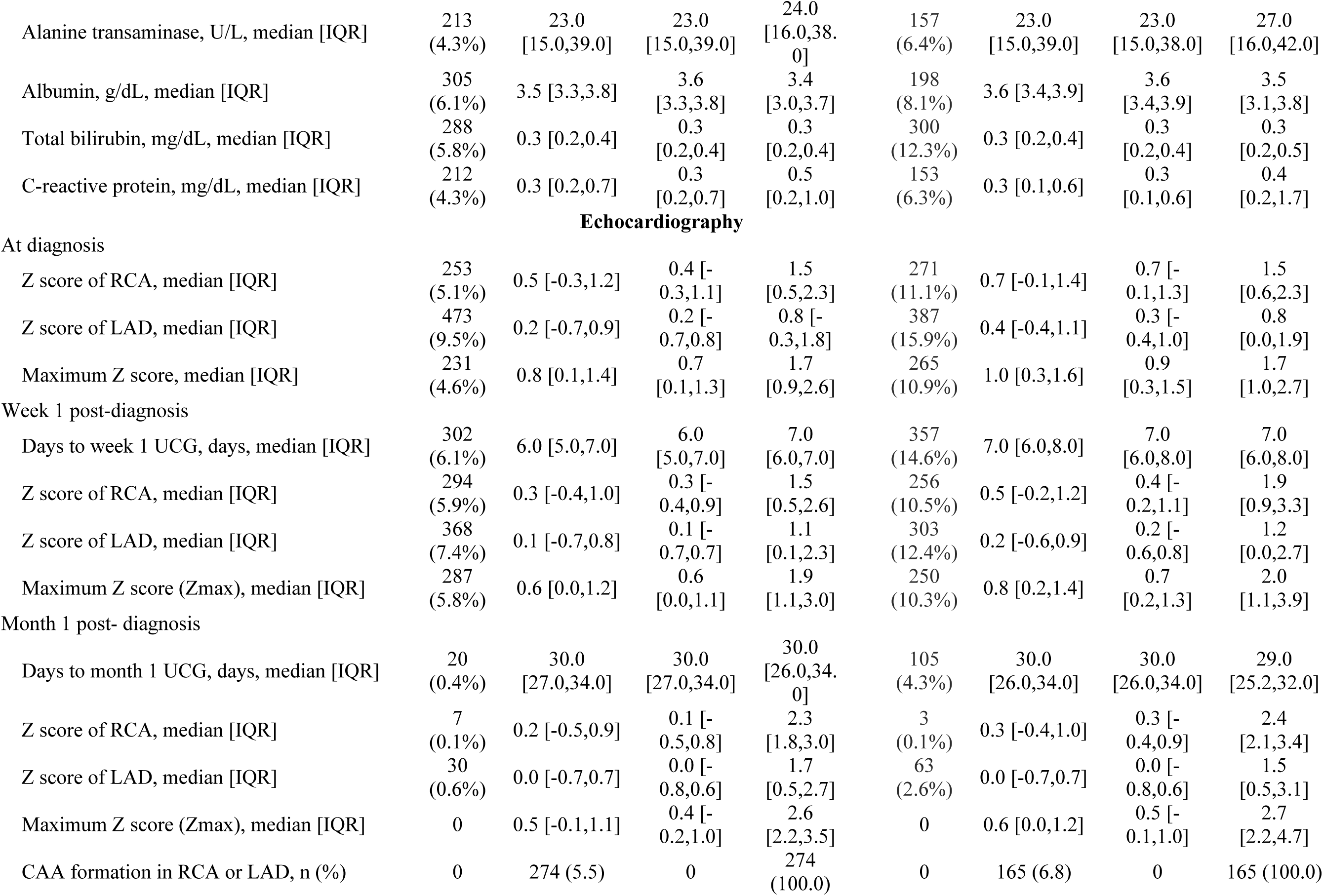
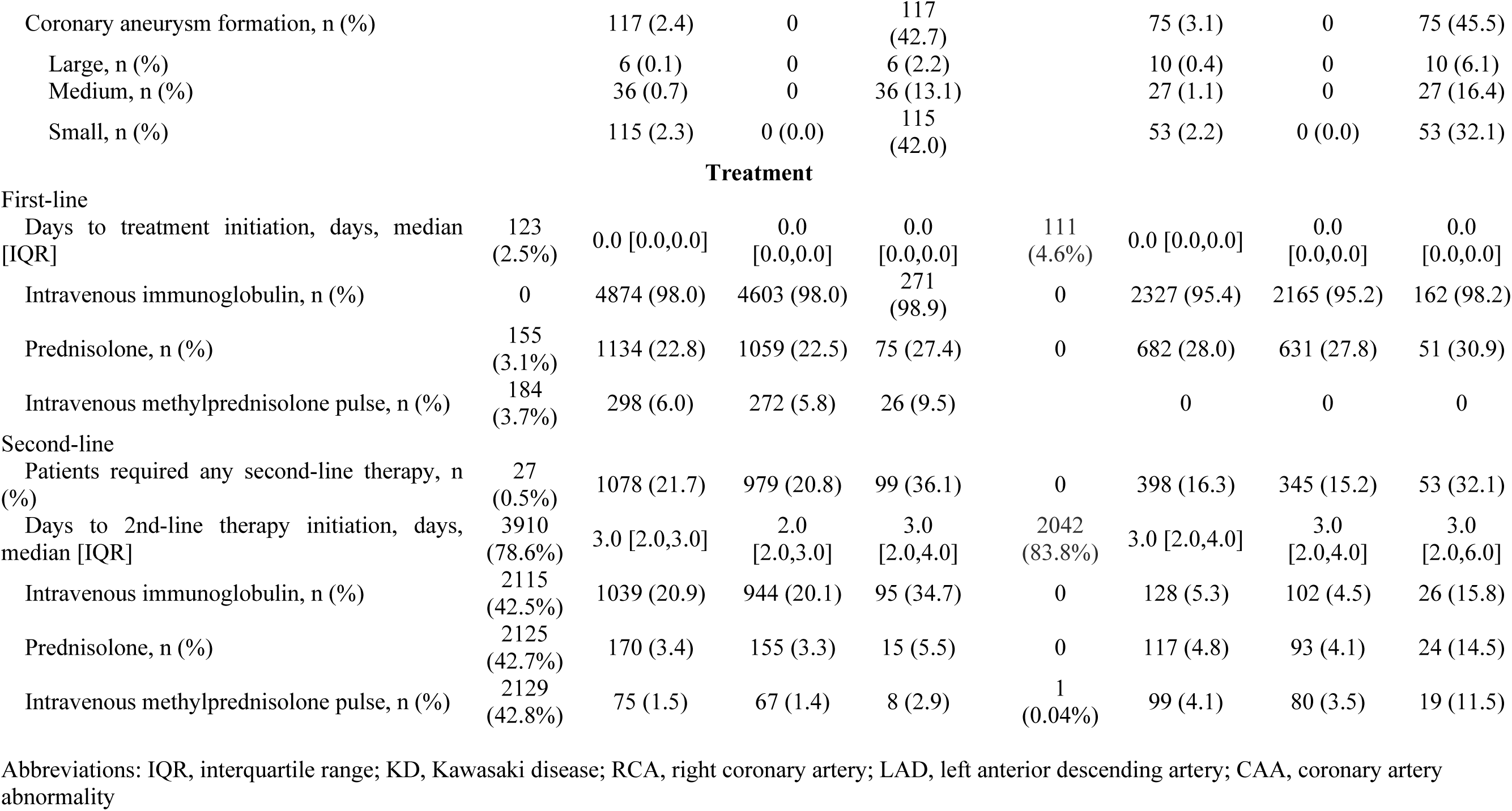
Patient characteristics.

Twenty-two models for predicting CAA formation at month 1 were developed and evaluated. Tables 3 and 4 show the AUC scores, and Figures 1 and 2 show the ROC curves of the main and exploratory analysis, respectively. Figures S1 and S2 show the calibration plots and the decision curves. The cutoff value maximizing specificity while maintaining a sensitivity of at least 0.95 in the PEACOCK cohort was applied to the Post-RAISE cohort. Table 5 summarizes the key performance metrics of the models in the Post-RAISE cohort, including sensitivity, specificity, PPV, NPV, proportion of potential avoidable echocardiograms, and number of false-negative cases obtained using this cutoff.

**Figure 1.**
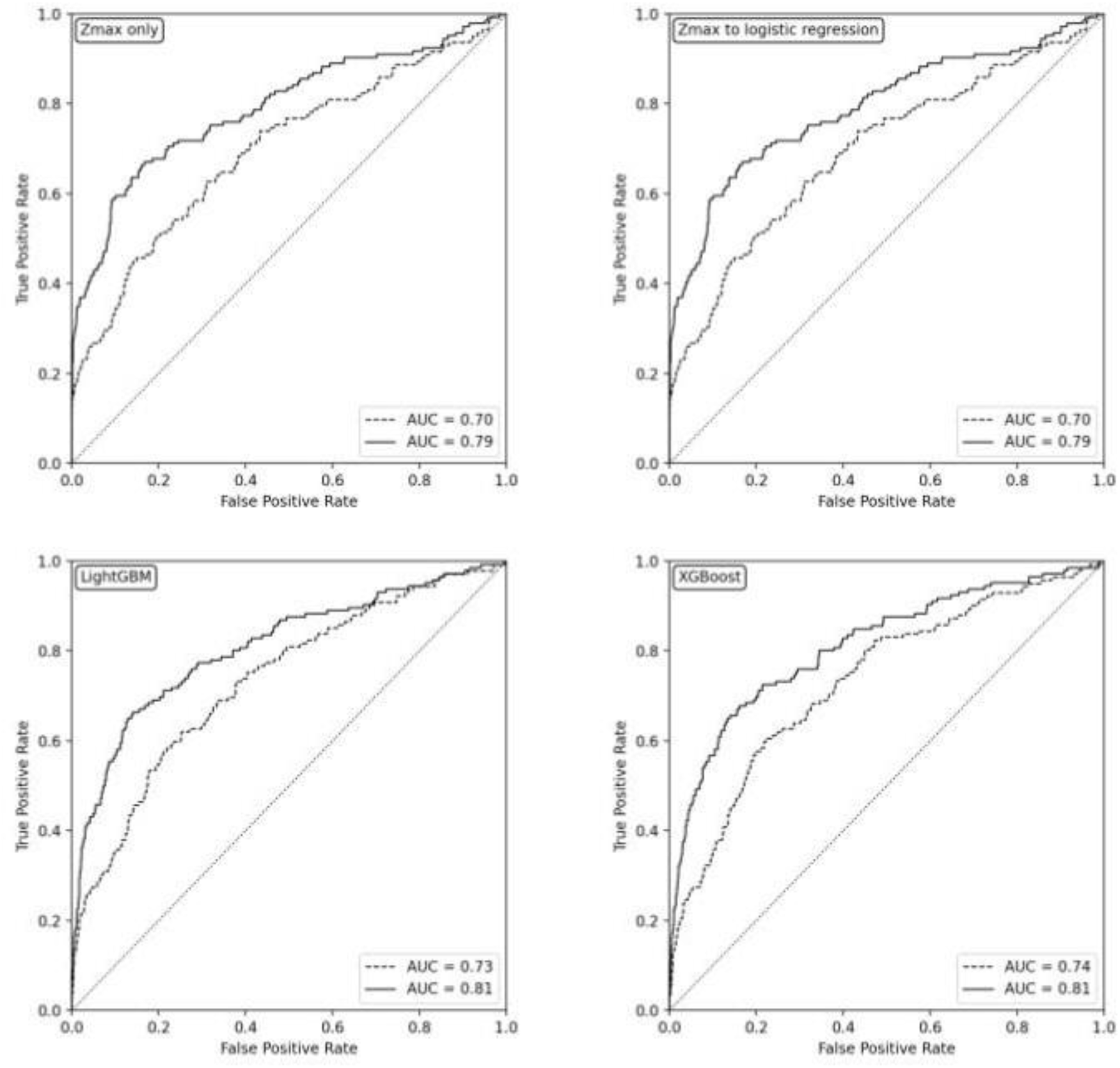
Receiver operating characteristic curves in the main analysis Each panel corresponds to the type of model used. The solid lines represent models built with data obtained up to week 1 post-diagnosis while the broken lines indicate models using data obtained at diagnosis only.

**Figure 2.**
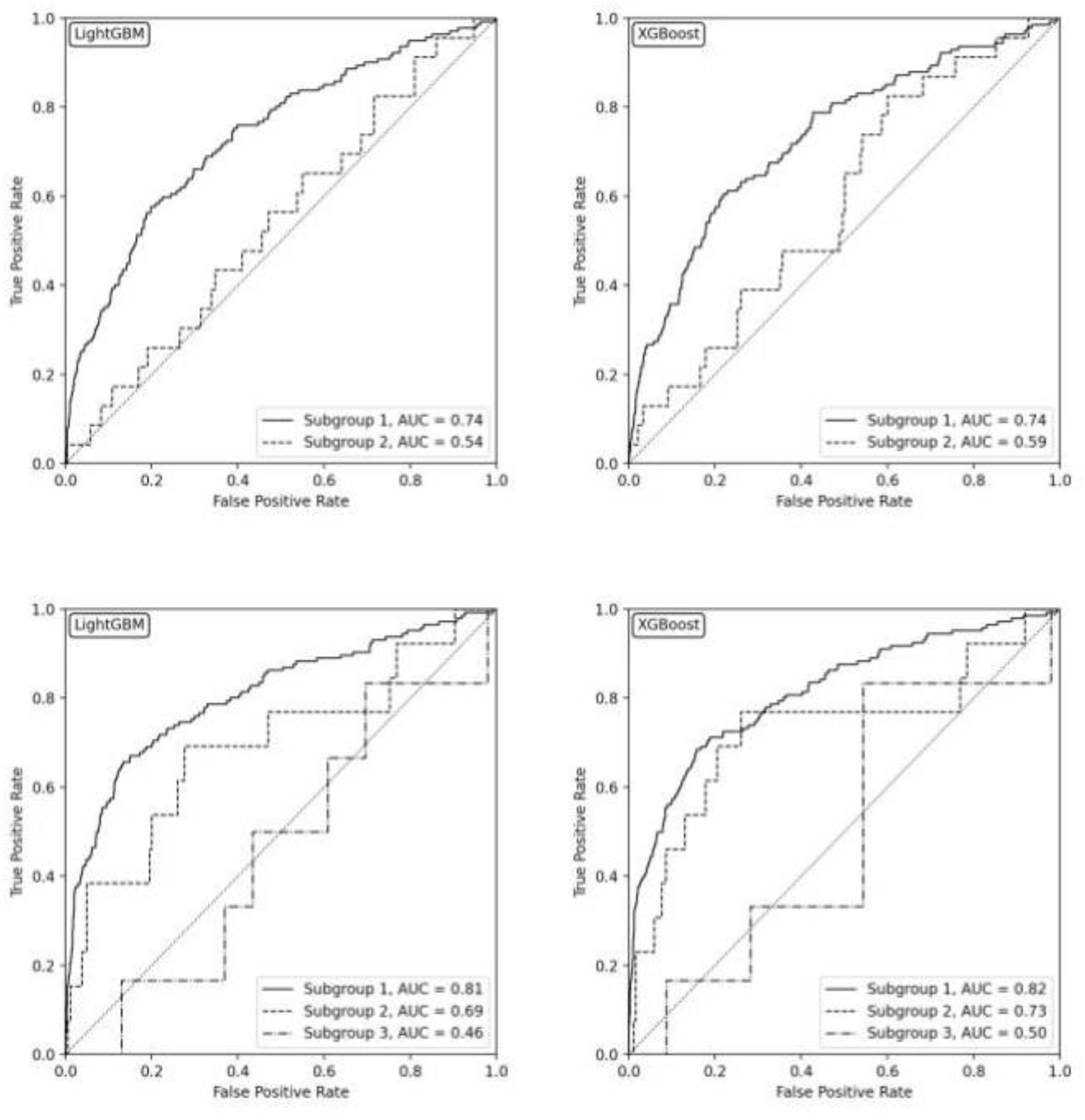
Receiver operating characteristic curves in the exploratory analyses The two upper panels show the results of analyses using data available at diagnosis while the two lower panels show the results of analyses using data obtained up to week 1 post-diagnosis. In the upper panels, the solid lines represent models built with data on patients with the Zmax value at diagnosis. The broken lines represent models built using data from patients without the Zmax value at diagnosis. In the lower panels, the solid lines represent models built with data from patients with the Zmax value at the week 1 assessment. The broken lines represent models built with data on patients without the Zmax value at week 1 but having the Zmax at diagnosis. The dash-and-dot lines represent models built with data on patients without any Zmax values.

**Table 3.**
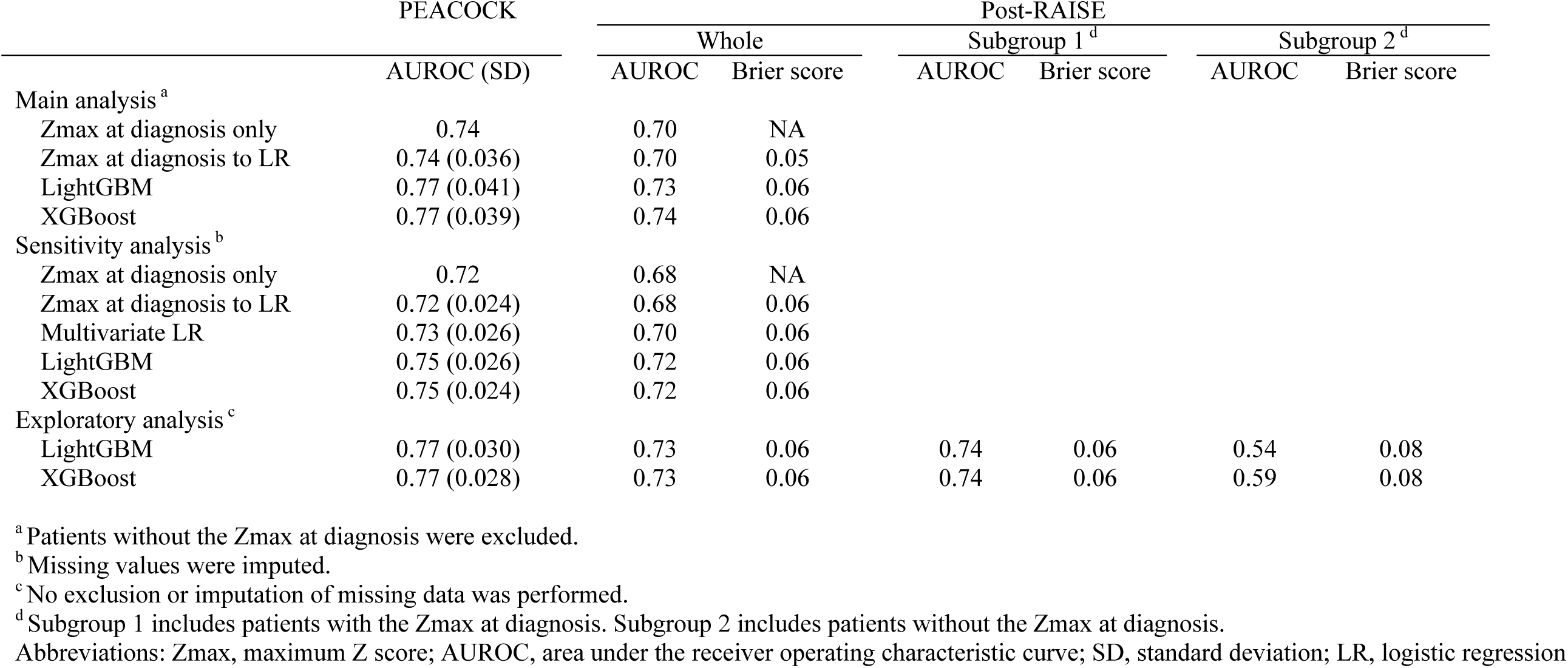
Model performance in each analytical method using data obtained at diagnosis only.

**Table 4.**
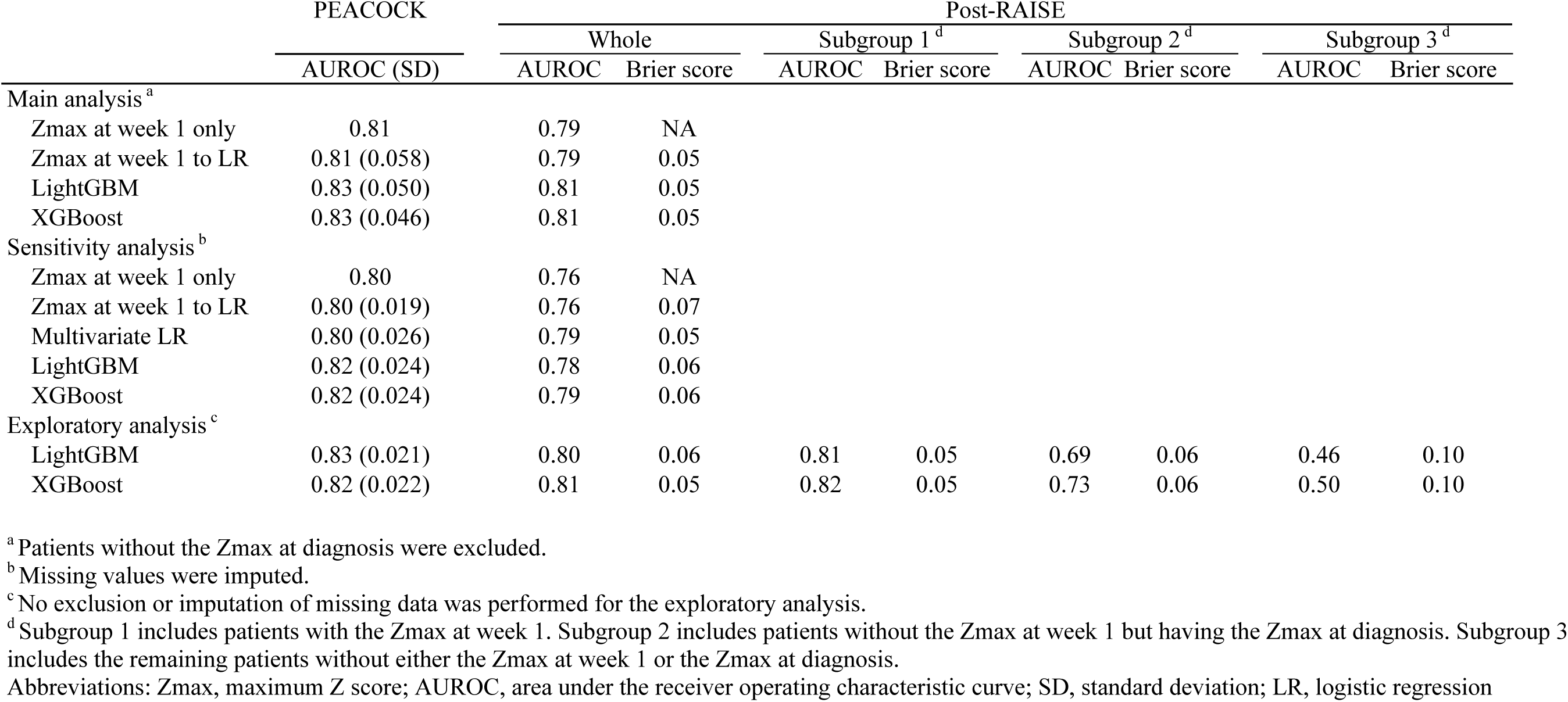
Model performance in each analytical method using data obtained at diagnosis and one week later.

**Table 5.** Statistical metrics, reduction in echocardiography need, and of false-negative cases.

|  | Sensitivity (%) | Specificity (%) | PPV (%) | NPV (%) | Potential echocardiogram reduction (%)<sup>a</sup> | Number of false negative cases (b) |  |  |  |  |
| --- | --- | --- | --- | --- | --- | --- | --- | --- | --- | --- |
|  |  |  |  |  |  | Total (%) | Dilation only | Small aneurysm | Medium aneurysm | Large aneurysm |
| Using data obtained at diagnosis only |  |  |  |  |  |  |  |  |  |  |
| Main analysis |  |  |  |  |  |  |  |  |  |  |
| Zmax at diagnosis only | 93.7% | 9.4% | 6.7% | 95.5% | 9.2% | 9 (0.4%) | 3 | 4 | 0 | 2 |
| Zmax at diagnosis to LR | 93.7% | 12.1% | 6.9% | 96.5% | 11.7% | 9 (0.4%) | 3 | 4 | 0 | 2 |
| LightGBM | 92.3% | 24.6% | 7.9% | 97.8% | 23.5% | 11 (0.5%) | 6 | 3 | 1 | 1 |
| XGBoost | 84.5% | 39.2% | 8.9% | 97.3% | 37.2% | 22 (1.0%) | 10 | 8 | 2 | 2 |
| Sensitivity analysis |  |  |  |  |  |  |  |  |  |  |
| Zmax at diagnosis only | 94.5% | 9.0% | 7.0% | 95.8% | 8.8% | 9 (0.4%) | 3 | 4 | 0 | 2 |
| Zmax at diagnosis to LR | 100.0% | 1.3% | 6.8% | 100.0% | 1.2% | NA | NA | NA | NA | NA |
| Multivariate LR | 95.8% | 11.1% | 7.3% | 97.3% | 10.6% | 7 (0.3%) | 4 | 2 | 0 | 1 |
| LightGBM | 90.3% | 30.7% | 8.6% | 97.8% | 29.3% | 16 (0.7%) | 9 | 5 | 1 | 1 |
| XGBoost | 82.4% | 46.0% | 10.0% | 97.3% | 44.1% | 29 (1.2%) | 15 | 10 | 2 | 2 |
| Using data obtained at both diagnosis and one week after |  |  |  |  |  |  |  |  |  |  |
| Main analysis |  |  |  |  |  |  |  |  |  |  |
| Zmax at diagnosis only | 92.5% | 19.2% | 7.6% | 97.3% | 18.4% | 11 (0.5%) | 4 | 6 | 1 | 0 |
| Zmax at diagnosis to LR | 91.1% | 23.3% | 7.8% | 97.3% | 22.3% | 13 (0.6%) | 4 | 8 | 1 | 0 |
| LightGBM | 93.2% | 28.9% | 8.6% | 98.3% | 27.4% | 10 (0.5%) | 7 | 3 | 0 | 0 |
| XGBoost | 84.9% | 54.0% | 11.7% | 98.0% | 51.4% | 22 (1.0%) | 11 | 9 | 1 | 1 |
| Sensitivity analysis |  |  |  |  |  |  |  |  |  |  |
| Zmax at diagnosis only | 92.7% | 17.6% | 7.5% | 97.1% | 16.9% | 12 (0.5%) | 4 | 7 | 1 | 0 |
| Zmax at diagnosis to LR | 92.1% | 21.7% | 7.9% | 97.4% | 20.8% | 13 (0.5%) | 4 | 8 | 1 | 0 |
| Multivariate LR | 93.3% | 18.9% | 7.7% | 97.5% | 18.1% | 11 (0.5%) | 5 | 6 | 0 | 0 |
| LightGBM | 90.9% | 26.2% | 8.2% | 97.5% | 25.1% | 15 (0.6%) | 7 | 7 | 1 | 0 |
| XGBoost | 79.4% | 57.6% | 12.0% | 97.5% | 55.1% | 34 (1.4%) | 20 | 11 | 2 | 1 |

A cutoff value maximizing specificity while maintaining sensitivity $\geq 0.95$ in the PEACOCK cohort was identified and applied to the Post-RAISE cohort. Various performance metrics and the number of false-negative cases are shown.
<sup>a</sup>Proportion of patients not predicted to develop coronary artery abnormalities at month 1.
<sup>b</sup>Number of patients with coronary artery abnormality development contrary to prediction. Coronary artery abnormality was defined by the maximum Z score: dilation only, $\geq 2$ to $<2.5$ ; small aneurysm, $\geq 2.5$ to $<5$ ; medium aneurysm, $\geq 5$ to $<10$ ; large aneurysm, $\geq 10$ .
Abbreviations: Zmax, maximum Z score; LR, logistic regression; PPV, positive predictive value; NPV, negative predictive value.

Tables 3 and 4 show that the raw Zmax models using the Zmax obtained at diagnosis and week 1 post-diagnosis individually demonstrated moderate discriminatory performance (AUC: 0.70 and 0.79, respectively). Adding other variables improved discrimination by less than 0.03 even when the machine learning models were used. Sensitivity analysis using imputed datasets demonstrated a similar trend. Exploratory analysis revealed that the AUC decreased in the absence of the Zmax value at baseline and week 1 post-diagnosis. Table 5 shows that none of the models reduced the need for echocardiography at month 1 by more than 30% while maintaining the number of misdiagnoses of CAA to less than ten. More than half these cases involved coronary aneurysms (Zmax ≥ 2.5) rather than coronary artery dilatation (2 ≤ Zmax < 5).

## DISCUSSION

### Summary of findings

This study aimed to develop and validate models for predicting CAA development at month 1 post-diagnosis to determine the need for follow-up echocardiography. The results demonstrated that the Zmax value based on echocardiographic findings at week 1 post-diagnosis had modest predictive value. Adding other clinical, laboratory, and treatment-related variables only marginally improved the prediction.

### Clinical implications

Although the models using the data obtained up to one week after diagnosis had a high NPV, their performance was still insufficient for clinical application. None of the models reduced the need for follow-up echocardiography at month 1 by more than 30% without overlooking at least ten patients with CAA development. Even the most complex models, designed with a low threshold to optimize sensitivity, failed to minimize the risk of overlooking CAA development to a clinically acceptable degree while reducing the need for echocardiography. Avoiding unnecessary echocardiography is beneficial to patients, their family, healthcare providers, and payers, but these benefits must be weighed carefully against the danger of overlooking patients requiring further evaluation and treatment.^31^ Therefore, the models may not yet be suitable for use in clinical practice. Until superior, predictive parameters or the optimal timing for interim echocardiography is determined, follow-up echocardiography at month 1 should remain the standard of care.

### Importance of the Zmax

Our study’s most striking finding was that the Zmax at week 1, by itself, was a powerful predictor of CAA at month 1 and that even complex machine learning models incorporating 28 additional variables achieved only marginal improvement over its predictive performance.

Additionally, the exploratory analysis indicated that excluding the Zmax from the multivariable machine learning models markedly reduced the AUC. The predictive utility of early Zmax values, in contrast to the limited predictive value of the other variables, warrants deeper consideration.

Various reasons explain the predictive value of the Zmax at week 1. First, the value is not merely a measurement of the coronary artery diameter at a single point in time but is also an integrated, summary measure of the cumulative effect of the initial disease activity, the host’s inflammatory response, and crucially, the individual’s biological response to the initial therapy, such as IVIG. It is possible that as a dynamic, echocardiographic parameter, the Zmax reflects much of the predictive information contained within baseline biomarkers, such as the CRP value and neutrophil count. Second, attempting to predict the outcome at month 1 using static, pre-treatment data is challenging owing to the significant influence of the therapeutic response. This may explain why our models, despite their sophistication, failed to find a strong predictive signal from the numerous baseline variables. This finding further suggested that to predict the prognosis of KD, dynamic parameters reflecting the therapeutic response may be more valuable than a large set of static baseline predictors.

### Improving prediction

Efforts to identify better predictors, including novel biomarkers, are ongoing. In an umbrella review of meta-analyses, Kim et al. reported that non-coding RNAs and the prognostic nutritional index may predict CAA development.^32^ Zheng et al. suggested that α-1-antitrypsin short peptide and guanine nucleotide-binding protein G(i) subunit alpha-2 also have the potential to predict CAA development.^33^ Identifying causal factors of Kawasaki disease will undoubtedly enhance predictive accuracy, and factors that improve prediction may, in turn, provide insight into the underlying cause. Further research is needed to discover more accurate and practical biomarkers.

In addition to novel predictors, re-evaluating the timing of interim echocardiography may help reduce unnecessary imaging tests. Echocardiography is typically performed at baseline, week 1 or 2 (interim assessment), and again at month 1 to assess for CAA in patients without complications. In our cohorts, the interim assessments were conducted four to nine days after diagnosis. Performing interim echocardiography later in the course may reduce false negatives and increase true negatives. De Ferranti et al. reported that among 464 patients who underwent echocardiography at baseline and week 2 (days 7–21) after fever onset, 456 (98.3%) maintained Zmax < 2 at weeks 3–9.^7^ Thus, delaying interim echocardiography in their cohort may have contributed to a greater reduction of 55.5% (464/844 patients) of the need for follow-up echocardiography at month 1 while maintaining a high NPV of 98.3%. Despite the potential improvement in prediction, delaying the interim assessment may theoretically lead to delayed treatment in patients with CAA. Therefore, determining the optimal timing for interim echocardiography remains an important area for future investigation.

### Limitations

This study has several limitations. First, overlooking patients with a Zmax slightly higher than 2 at month 1 may have only marginal clinical significance, as most cases of minor CAA resolve spontaneously.^14^ Distinguishing between transient and persistent abnormalities and identifying patients requiring intervention were not feasible owing to the unavailability of follow-up data beyond month 1 post-diagnosis. Second, the findings of this study may not be generalizable; although the study analyzed two large, prospectively collected datasets from multiple institutions in Japan, the findings may not apply to other ethnic groups or healthcare systems. Therefore, external validation by studies enrolling a more diverse sample population is warranted. Third, this study did not assess cost-effectiveness or consider the perspectives of patients and caregivers, both of which are important for implementing changes to standard care.^7^ Nonetheless, this study provided added value by analyzing two large cohorts, which enabled external validation, and by incorporating machine learning models with traditional algorithms.

## CONCLUSIONS

Using two large Japanese cohorts, this study created and externally validated 22 models for predicting CAA development at month 1 post-KD diagnosis. The Zmax obtained at week 1 post-diagnosis performed comparably with more complex models employing 29 variables.

However, none of the models achieved the accuracy necessary to reduce the need for echocardiography while limiting the risk of overlooking CAA development. Until more robust predictors are discovered, follow-up echocardiography at month 1 should remain the standard practice as recommended by the current, clinical guidelines.

## Data Availability

Anonymized individual participant data are kept confidential and will not be made available.

## Acknowledgments

We thank James R. Valera for editorial assistance; Kazutaka Yoshinaga and Ryo Ueno for valuable advice on study design and statistical analysis; and Masako Tomotsune and the staff of the Clinical Research Support Center at Tokyo Metropolitan Children’s Medical Center for assistance with data management.

## Supplemental Material

Supplementary Methods. Undersampling with bagging

Table S1. Predictors selected for model development

Table S2. Comparison of patient characteristics of the included and excluded patients derived from the original PEACOCK cohort

Table S3. Comparison of patient characteristics of the included and excluded patients derived from the original Post-RAISE cohort

Figure S1. Calibration plots and decision curves for the main analysis using data obtained at diagnosis

Figure S2. Calibration plots and decision curves for the main analysis using data obtained at both diagnosis and one week after

## Non-standard Abbreviations and Acronyms

KD: Kawasaki disease
CAA: coronary artery abnormality
PEACOCK: the Prospective Study on Efficacy of Acute Treatment in a Multicenter Cohort of Children with Kawasaki Disease
Post-RAISE: the Prospective Observational study on STRAtified treatment with Immunoglobulin plus Steroid Efficacy for Kawasaki disease
IVIG: intravenous immunoglobulin
IVMP: intravenous methylprednisolone pulse therapy
RCA: proximal right coronary artery
LAD: proximal left anterior descending artery
CRP: C-reactive protein
ROC: receiver operating characteristic
AUC: area under the curve
PPV: positive predictive value
NPV: negative predictive value

